# Comparison of mental health service activity before and shortly after UK social distancing responses to the COVID-19 pandemic: February-March 2020

**DOI:** 10.1101/2020.09.26.20202150

**Authors:** Robert Stewart, Evangelia Martin, Ioannis Bakolis, Matthew Broadbent, Nicola Byrne, Sabine Landau

## Abstract

This study sought to provide an early description of mental health service activity before and after national implementation of social distancing for COVID-19. A time series analysis was carried out of daily service-level activity on data from a large mental healthcare provider in southeast London, from 01.02.2020 to 31.03.2020, comparing activity before and after 16.03.2020: i) inpatient admissions, discharges and numbers, ii) contact numbers and daily caseloads (Liaison, Home Treatment Teams, Community Mental Health Teams); iii) numbers of deaths for past and present patients. Daily face-to-face contact numbers fell for liaison, home treatment and community services with incomplete compensatory rises in non-face-to-face contacts. Daily caseloads fell for all services, apart from working age and child/adolescent community teams. Inpatient numbers fell 13.6% after 16^th^ March, and daily numbers of deaths increased by 61.8%.

## INTRODUCTION

The first wave of the COVID-19 pandemic had an impact across many healthcare sectors: not only because of the direct effects of the virus itself on communities and healthcare staff, but also arising from the national public health policies enacted to reduce spread. Mental healthcare faced a range of challenges including the heightened vulnerability of its patient populations (e.g. through cardiovascular and respiratory disorders), already-reduced life-expectancies (1), and frequently described problems accessing healthcare (2;3). In addition, services had to be radically reconfigured to cope with suspected or confirmed COVID-19 infections in inpatient and outpatient settings, staff sickness or self-isolation, the need to minimise face-to-face contacts, and the need to accommodate increasing pressures on acute medical care from cases of viral pneumonia. These in turn were accompanied by the as yet unknown impacts of social distancing on already isolated or otherwise vulnerable populations, and of challenged national economies on already impoverished and disadvantaged communities. There therefore continues to be a pressing need for research (4). Taking advantage of a mental healthcare data platform that receives 24-hourly updates from its source electronic records, we sought to describe daily activity in key services for the months of February and March 2020 and to quantify statistically the early changes observed.

## METHODS

The National Institute for Health Research (NIHR) Biomedical Research Centre (BRC) Case Register at the South London and Maudsley NHS Foundation Trust (SLaM) has been described previously (5;6). In summary, SLaM is one of Europe’s largest mental healthcare providers, serving a geographic catchment of four south London boroughs (Croydon, Lambeth, Lewisham, Southwark) with a population of around 1.2 million residents. SLaM has used a fully electronic health record (EHR) across all its services since 2006, and the NIHR BRC Case Register was set up in 2008, providing researcher access to de-identified data from SLaM’s EHR via the Clinical Record Interactive Search (CRIS) platform and within a robust, patient-led security model and governance framework (7). CRIS has been extensively developed over the last 10 years with a range of external data linkages and natural language processing resources (6). Of relevance to the work presented here, CRIS is updated from SLaM’s EHR every 24 hours and thus provides relatively ‘real-time’ data, although prior to the COVID-19 pandemic had mostly been used to support historic cohort analyses. SLaM’s EHR is itself immediately updated every time an entry is made, which include date-stamped fields indicating patient contacts (‘events’) and those indicating acceptance of a referral, a discharge from a given service (or SLaM care more generally), including admissions to and discharges from inpatient care. Mortality in the complete EHR (i.e. all SLaM patients with records, past or present) is ascertained weekly through automated checks of National Health Service (NHS) numbers (a unique identifier used in all UK health services) against a national spine. CRIS has supported over 200 peer reviewed publications to date. CRIS has received approval as a data source for secondary analyses (Oxford Research Ethics Committee C, reference 18/SC/0372).

Activity and caseload data were extracted via CRIS and enumerated for every day from 1^st^ February 2020 to 31^st^ March 2020. For inpatient care, the following were calculated: number of new admissions from the community, number of new discharges from inpatient care to the community, number of current inpatients. In addition, numbers of inpatients classified as on leave were calculated for illustration but not analysis. For other selected services, daily caseloads were calculated by ascertaining patients who were receiving active care from a given service on a given day, based on the date a referral to that service was recorded as accepted to the point a discharge was made from that service. Daily contact numbers were ascertained from recorded ‘events’ (i.e. standard case note entries) for that service and were divided into the following groups according to structured compulsory meta-data fields for that event in the EHR: i) face-to-face contacts attended; ii) non-face-to-face contacts attended; iii) appointments cancelled or not attended (‘DNA’); iv) contacts listed as ‘other’. Non-face-to-face contacts included those recorded as being made by email, fax, mail, phone, online, or video link. This fourth group was investigated with manual inspection and was found to comprise a miscellaneous collection of contacts, including those with other staff members, other services (e.g. social care) or with patients’ friends/family. The following SLaM services were chosen for description and comparison, on the basis that they represented the largest and most strategically important groupings:

1. Inpatient care.
2. Liaison Mental Health services. SLaM provides liaison services to four Acute Trusts on five hospital sites: King’s College Hospital, St Thomas’ Hospital, Guy’s Hospital, University Hospital Lewisham, and Croydon University Hospital. Data from all child, working age and older adult liaison teams were combined for description here.
3. Working age Home Treatment Team (HTT) services.
4. Older adults HTT services.
5. Working age Community Mental Health Team (CMHT) services.
6. Child and Adolescent Mental Health (CAMH) community services.
7. Older adults CMHT services.

Finally, mortality data (number of deaths for all patients with records) were extracted. Caseload and contact data were extracted on 2^nd^ April 2020; because of delays in registrations and incorporation of this information coming in the Case Register, number of deaths were extracted for the time period of interest on 23^rd^ April 2020.

The primary objective was to present descriptive data, which were displayed graphically (Mon-Sun for inpatient, liaison and HTT services; Mon-Fri for CMHT and CAMH services). In addition, caseloads and core activity (face-to-face and non-face-to-face contacts) were formally compared across the period of interest. In this respect, 16^th^ March was chosen as an index date, being the date on which the national self-isolation strategy was announced (https://www.gov.uk/government/speeches/pm-statement-on-coronavirus-16-march-2020). Activity levels before and after that date were therefore compared. Because of the heterogeneity in levels of service delivery at weekends, statistical comparisons considered the 40 weekday observations only (between the 3^rd^ of February and the 27^th^ March inclusive) for all measures. All variables represented counts of some type and were modelled using a negative binomial regression model to allow for overdispersion where this was indicated. To account for systematic trend over time, and also systematic weekly patterns the models included fixed effects of week (8 levels) and weekday (5 levels) factors. More complex modelling could not be supported by the relatively small sample size of observations to date. The negative binomial model assumed that the daily counts were statistically independent.

Time series data are often thought to display extra autocorrelations due to unaccounted short-term effects. To account for these, a sensitivity analysis that fitted an autoregressive correlation structure (with one autocorrelation parameter) to the daily error terms within a week was carried out using generalised estimating equations (GEE). Observations from different weeks were still assumed independent. GEE with a negative binomial distribution requires provision of the overdispersion parameter. This parameter was set to the value estimated in the negative binomial regression, provided the parameter had tested statistically significant at the 5% level. However, many of these models fitted a negative autocorrelation parameter, which may indicate unaccounted systematic trend rather than error dependencies. Either way, as will be described, inferential results were little affected by relaxing the independence assumption.

## RESULTS

Daily counts for each service are graphically displayed in Figures 1-14, and comparisons before and after the 16^th^ March are summarised in Table 1. For inpatient care, there was a decline in admissions and an increase in discharges (Figure 1), with an estimated 14% reduction in inpatient numbers after 16^th^ March (Figure 2, Table 1). For all other services, there was a common pattern of reduced face-to-face and increased non-face-to-face contacts per day, although estimated trends in combined contact numbers were always negative (Table 1). Liaison services (Figures 3-4) exhibited a substantial (estimated 36%) fall in total contacts, and a small (6%) but statistically significant overall reduction in daily caseload (Table 1). Both working age and older adult HTTs (Figures 5-8) showed reductions in total contacts (26% and 33% respectively) and daily caseloads (16% and 24% respectively), both more pronounced for the older adult services (Table 1). Considering community team services (Figures 9-14), all (working age, CAMH, older adult) showed reductions in total assessments (14%, 14% and 20% respectively), but only older adult daily caseloads fell (by 6%), while those for working age and CAMH services did not change significantly after the 16^th^ March (Table 1). Daily numbers of deaths are displayed in Figure 15 and showed a significant 62% increase after 16^th^ March (Table 1).

**Table 1:** Estimated incidence ratios (IRRs) comparing weekday mental health service activity (3^rd^ February to 27^th^ March 2020) before and after the national introduction of social distancing (16^th^ March)

| Measure (daily count) | Daily count (median, min-max) |  | IRR comparing before/after 16<sup>th</sup> March |  |  |  |
| --- | --- | --- | --- | --- | --- | --- |
|  | Before 16<sup>th</sup> March<br>(N=30 days) | 16<sup>th</sup> March and after<br>(N=10 days) | Estimate | 95% CI | p-value | p-value<br>from<br>sensitivity<br>analysis* |
| *Inpatient care* |  |  |  |  |  |  |
| Admissions | 12, 5-18 | 8.5, 5-12 | 0.687 | [0.542, 0.872] | 0.002 | <0.001 (-) |
| Discharges | 15, 7-26 | 23, 15-34 | 1.649 | [1.411, 1.927] | <0.001 | <0.001 (-) |
| Patient numbers | 738.5, 700-749 | 629.5, 576-701 | 0.864 | [0.840, 0.888] | <0.001 | <0.001 (+) |
| *Liaison services* |  |  |  |  |  |  |
| Face-to-face contacts | 150, 117-202 | 69, 53-112 | 0.478 | [0.441, 0.518] | <0.001 | <0.001 (-) |
| Non-face-to-face contacts | 35.5, 17-43 | 41, 36-67 | 1.328 | [1.183, 1.490] | <0.001 | <0.001 (-) |
| Total contacts | 184,156-243 | 119,91-155 | 0.640 | [0.610,0.680] | <0.001 | <0.001(-) |
| Caseloads | 2107, 2043-2141 | 1983, 1901-2051 | 0.942 | [0.927, 0.957] | <0.001 | <0.001 (+) |
| *Working age Home Treatment Team services* |  |  |  |  |  |  |
| Face-to-face contacts | 114, 97-163 | 66.5, 50-92 | 0.568 | [0.518, 0.622] | <0.001 | <0.001 (+) |
| Non-face-to-face contacts | 28, 19-39 | 41.5, 16-52 | 1.443 | [1.280, 1.626] | <0.001 | <0.001 (-) |
| Total contacts | 142,121-194 | 111, 92-121 | 0.740 | [0.688,0.795] | <0.001 | <0.001(-) |
| Caseloads | 183.5, 172-195 | 150.5, 138-180 | 0.839 | [0.793, 0.888] | <0.001 | <0.001 (+) |
| *Older adults Home Treatment Team services* |  |  |  |  |  |  |
| Face-to-face contacts | 26.5, 19-47 | 11, 6-31 | 0.462 | [0.380, 0.563] | <0.001 | <0.001 (-) |
| Non-face-to-face contacts | 1, 0-4 | 5.5, 0-10 | 3.645 | [2.327, 5.711] | <0.001 | <0.001 (-) |
| Total contacts | 28,21-48 | 18,14-31 | 0.669 | [0.562,0.782] | <0.001 | <0.001(-) |
| Caseloads | 38, 34-44 | 30, 21-38 | 0.764 | [0.671-0.870] | <0.001 | <0.001 (+) |
| *Working age Community Mental Health Team services* |  |  |  |  |  |  |
| Face-to-face contacts | 607, 462-714 | 253.5, 194-458 | 0.473 | [0.448, 0.498] | <0.001 | <0.001 (+) |
| Non-face-to-face contacts | 176, 132-218 | 385.5, 268-437 | 2.190 | [2.077, 2.308] | <0.001 | <0.001 (-) |
| Total contacts | 783, 651-893 | 682.5, 603-726 | 0.860 | [0.841, 0.890] | <0.001 | <0.001 (-) |
| Caseloads | 8841, 8779-8906 | 8886.5, 8867-8907 | 1.005 | [0.997, 1.012] | 0.226 | 0.380 (+) |
| *Child and Adolescent Mental Health services* |  |  |  |  |  |  |
| Face-to-face contacts | 209, 140-309 | 57, 34-122 | 0.279 | [0.251, 0.312] | <0.001 | <0.001 (-) |
| Non-face-to-face contacts | 82, 54-115 | 196, 145-220 | 2.331 | [2.158, 2.518] | <0.001 | <0.001 (-) |
| Total contacts | 299.5, 199-395 | 254, 211-287 | 0.855 | [0.814, 0.898] | <0.001 | <0.001 (-) |
| Caseloads | 6239.5, 6184-6312 | 6294, 6254-6322 | 1.008 | [0.999, 1.017] | 0.078 | 0.158 (+) |
| *Older adults Community Mental Health Team services* |  |  |  |  |  |  |
| Face-to-face contacts | 97, 58-131 | 34, 13-59 | 0.361 | [0.317, 0.411] | <0.001 | <0.001 (-) |
| Non-face-to-face contacts | 58, 41-87 | 85.5, 64-156 | 1.490 | [1.300, 1.708] | <0.001 | <0.001 (-) |
| Total contacts | 155.5, 110-203 | 119, 84-203 | 0.800 | [0.726, 0.882] | <0.001 | <0.001 (-) |
| Caseloads | 2237, 2177-2266 | 2094.5, 2015-2169 | 0.941 | [0.926, 0.955] | <0.001 | <0.001 (+) |
| Total deaths | 14.5, 7-21 | 21.5, 11-36 | 1.618 | [1.372, 1.908] | <0.001 | <0.001 (-) |

\*Allowing for an autoregressive correlation structure between weekdays within weeks.
(+) Fitting a positive residual correlation between adjacent days
(-) Fitting a negative residual correlation between adjacent days

**Figure 1:**
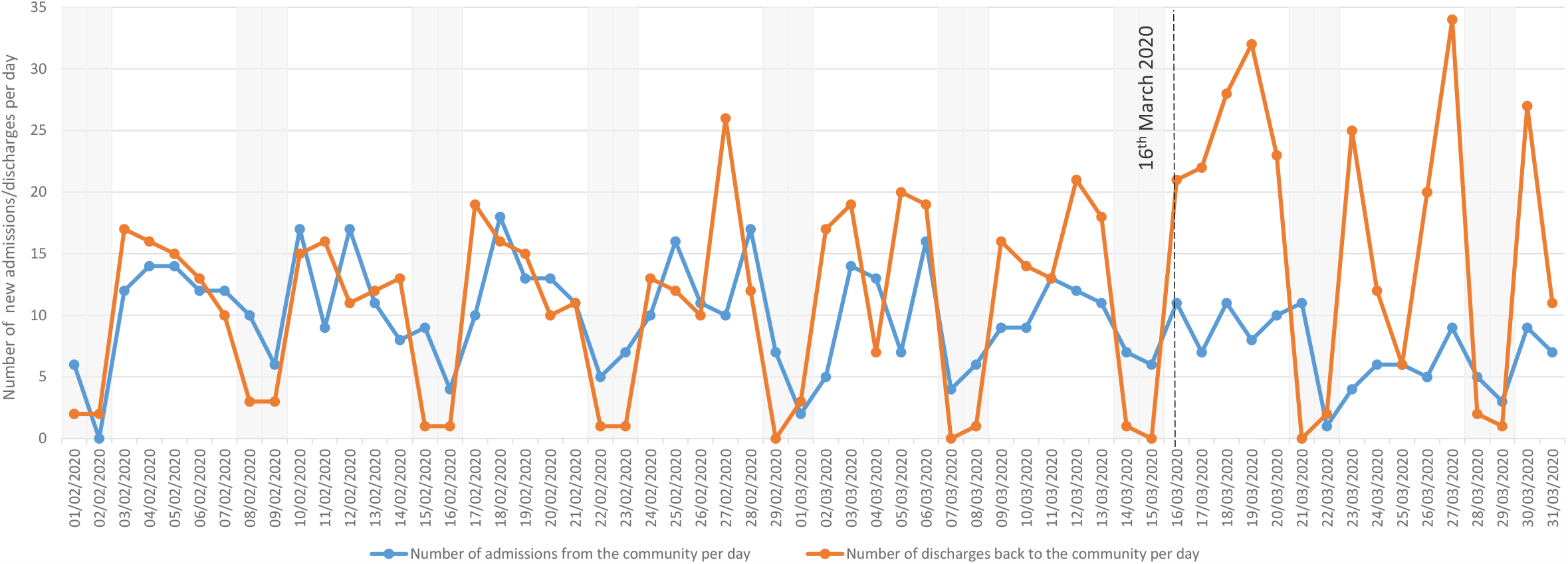
Numbers of new mental health inpatient admissions/discharges (daily; February-March 2020)

**Figure 2:**
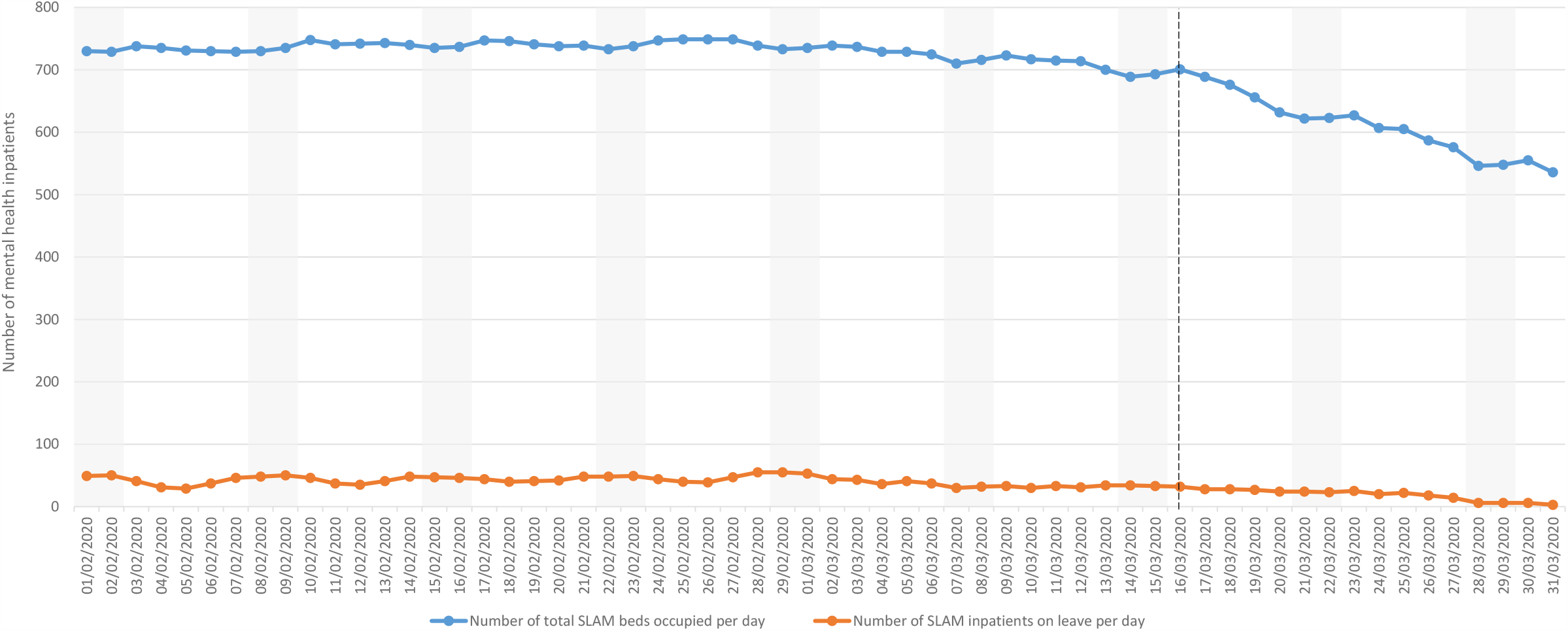
Total numbers of mental health inpatients (daily; February-March 2020)

**Figure 3:**
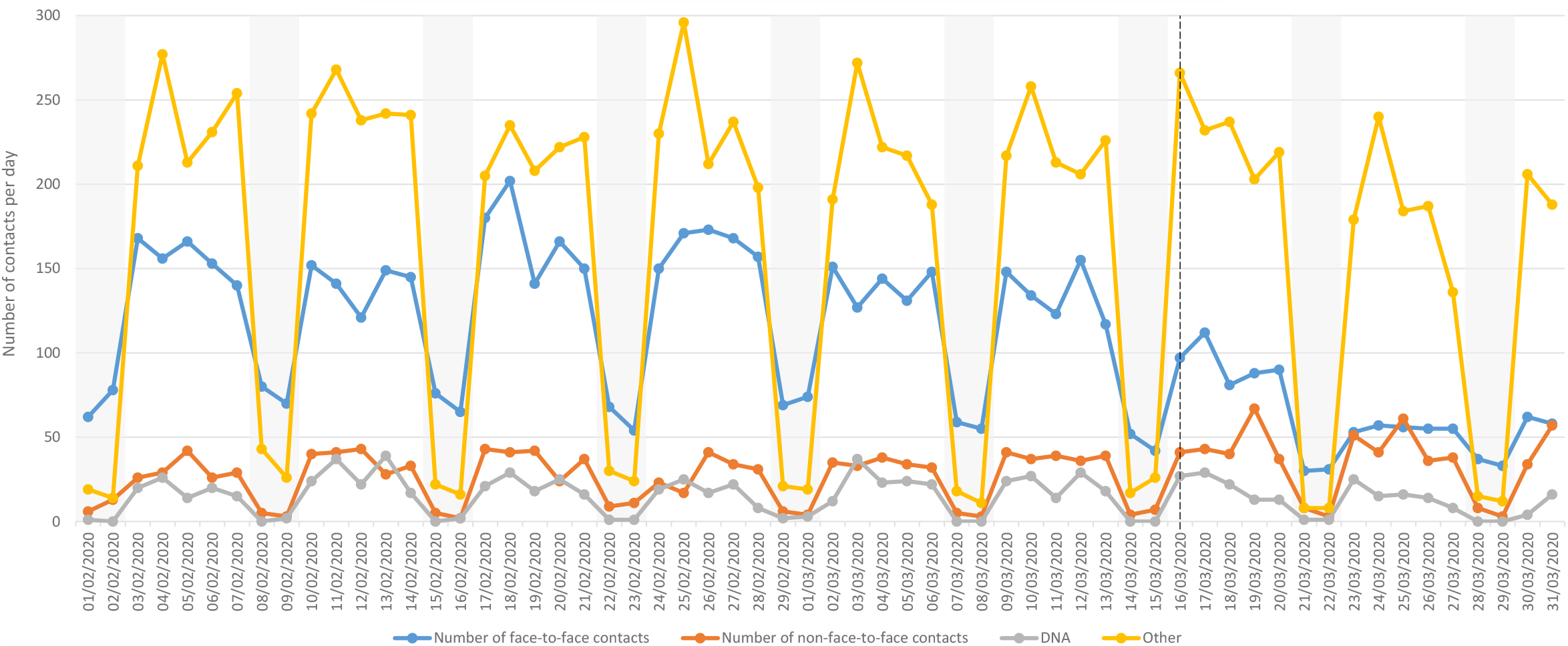
Liaison Service contacts by contact type (daily; February-March 2020)

**Figure 4:**
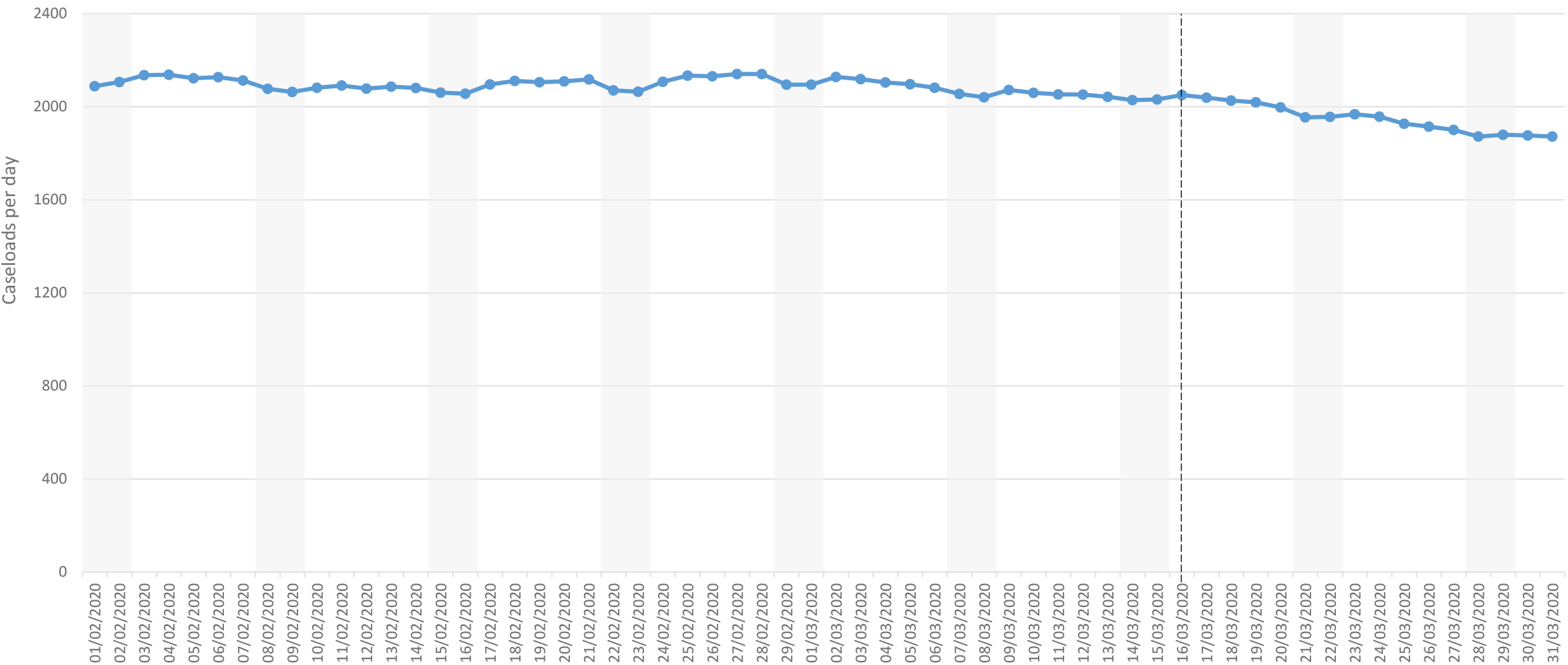
Liaison Service active caseloads (daily; February-March 2020)

**Figure 5:**
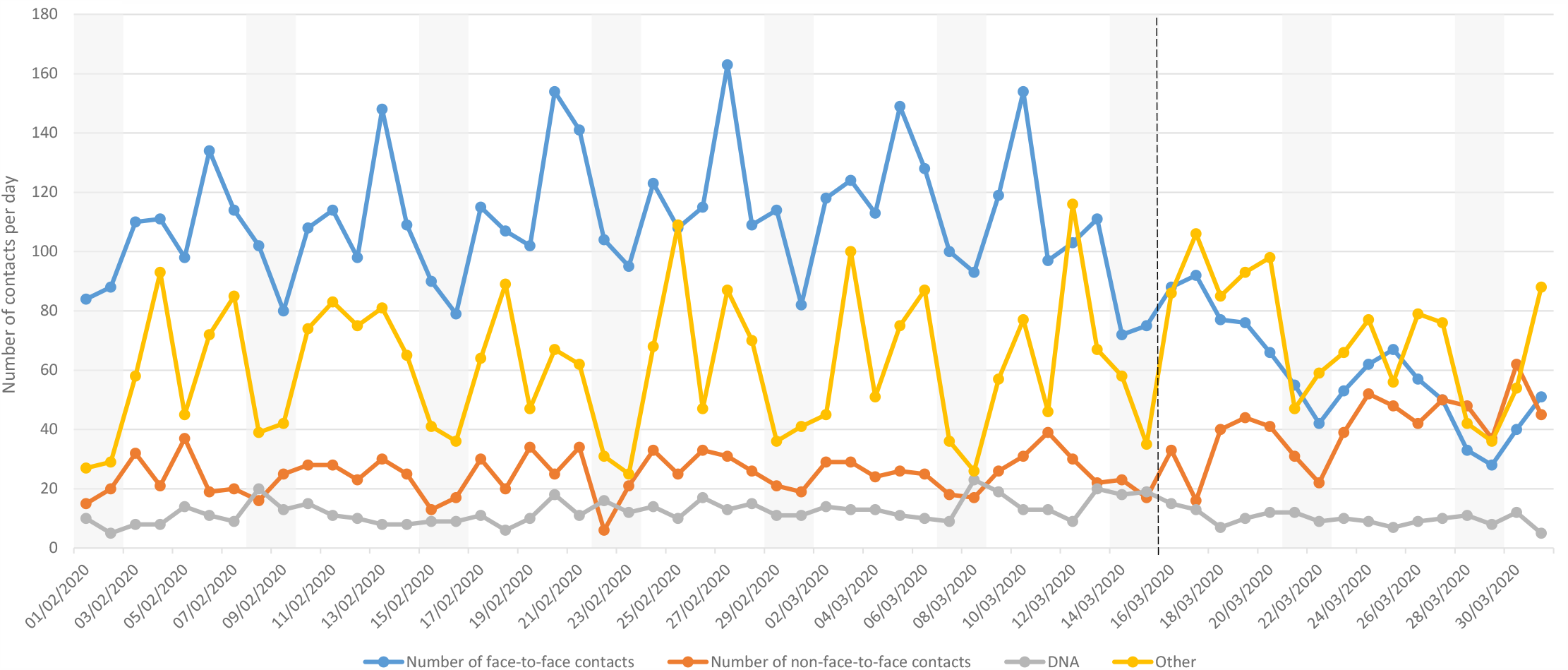
Working age adult Home Treatment Team community contacts by contact type (daily; February-March 2020)

**Figure 6:**
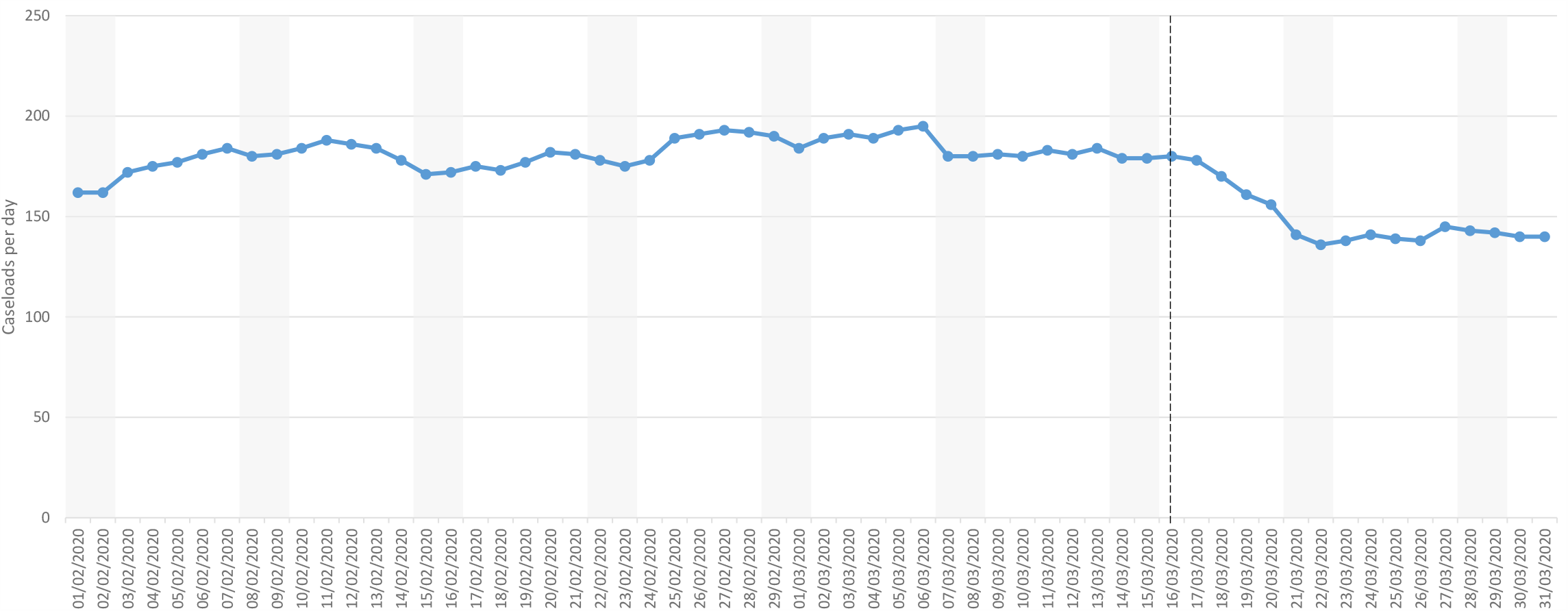
Working age adult Home Treatment Team active caseloads (daily; February-March 2020)

**Figure 7:**
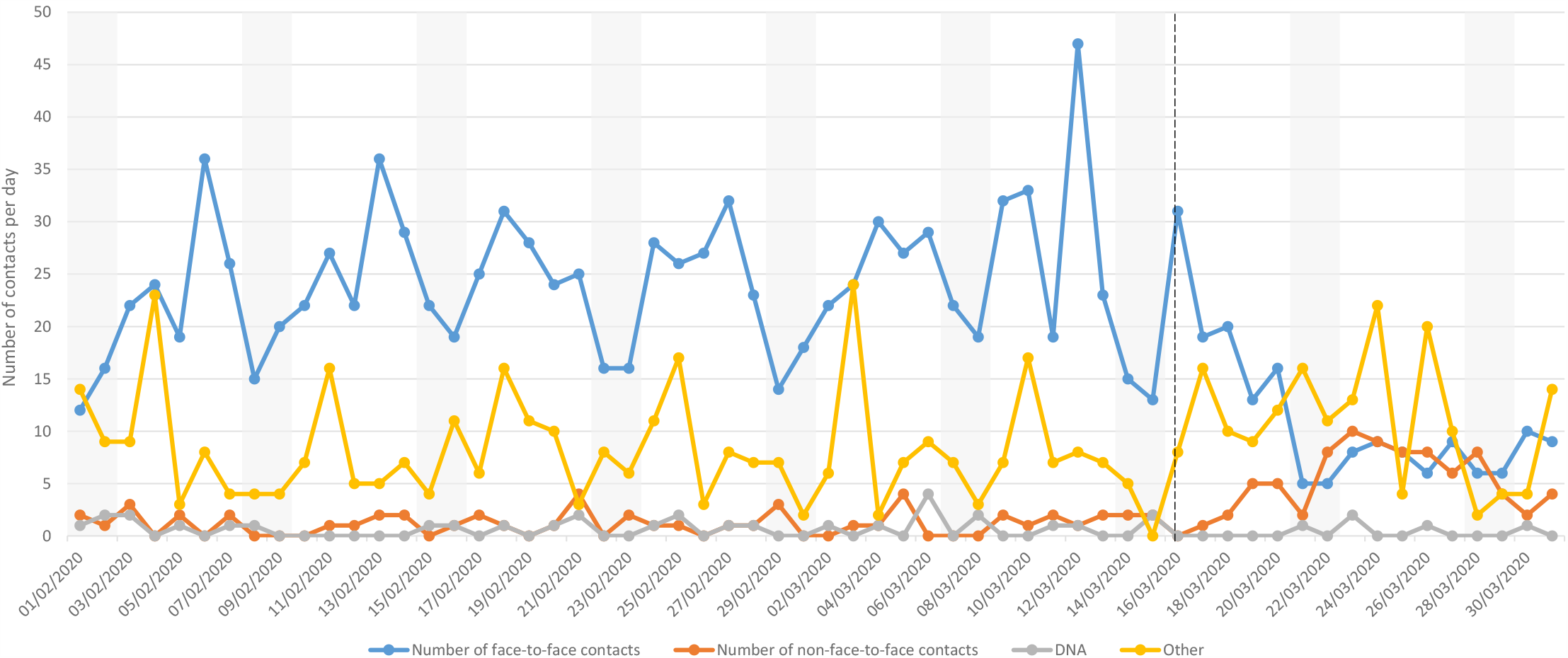
Older adult Home Treatment Team contacts by contact type (daily; February-March 2020)

**Figure 8:**
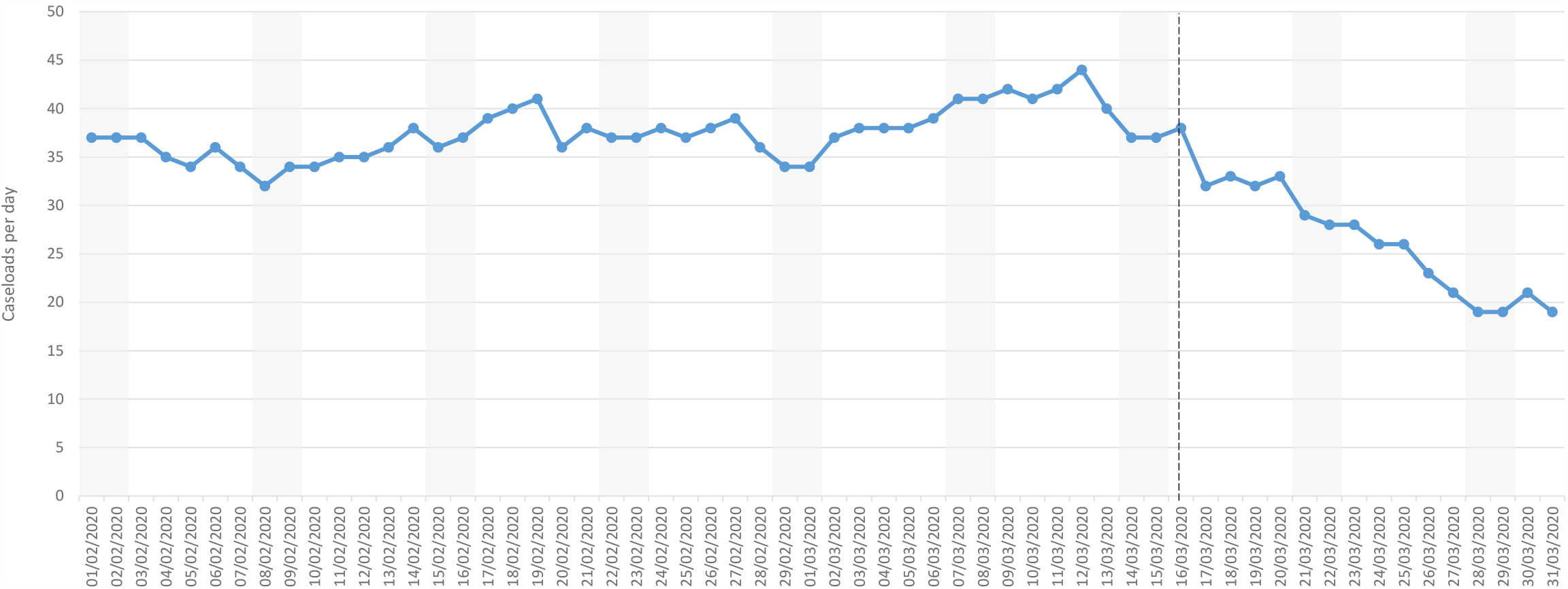
Older adult Home Treatment Team active caseloads (daily; February-March 2020)

**Figure 9:**
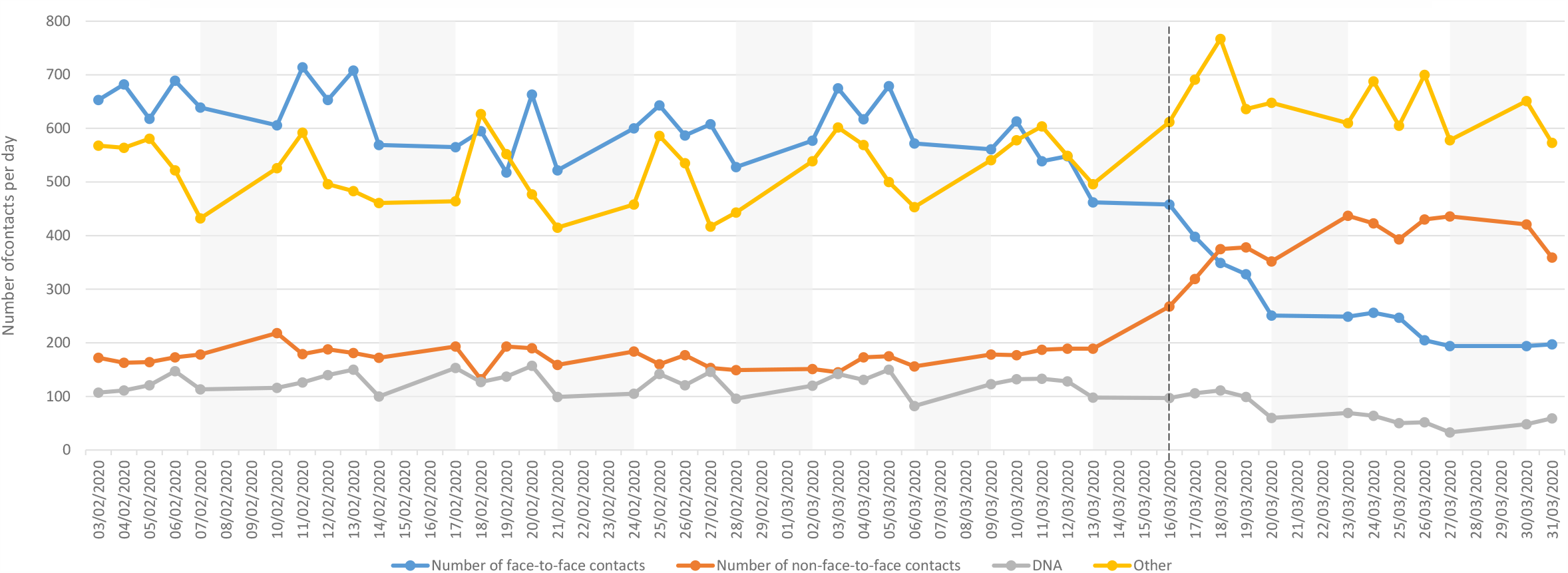
Working age adult Community Mental Health Team contacts by contact type (daily; Monday-Friday; February-March 2020)

**Figure 10:**
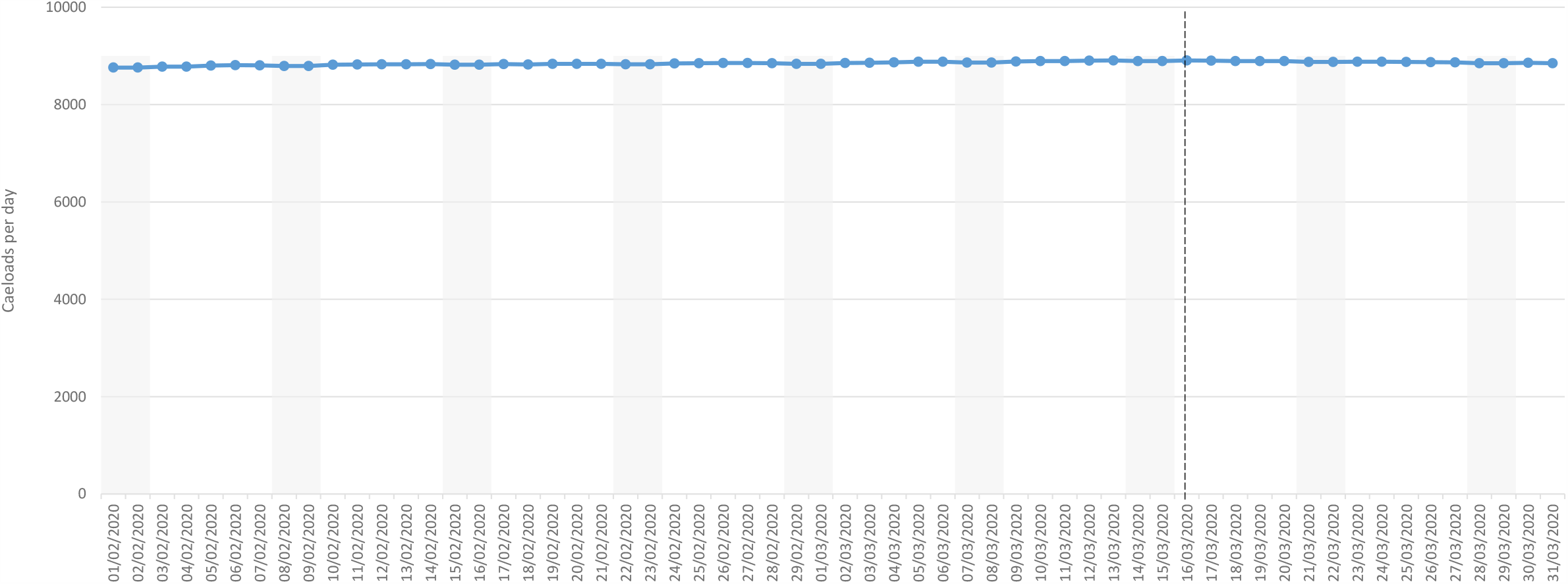
Working age adult Community Mental Health Team active caseloads (daily; February - March 2020)

**Figure 11:**
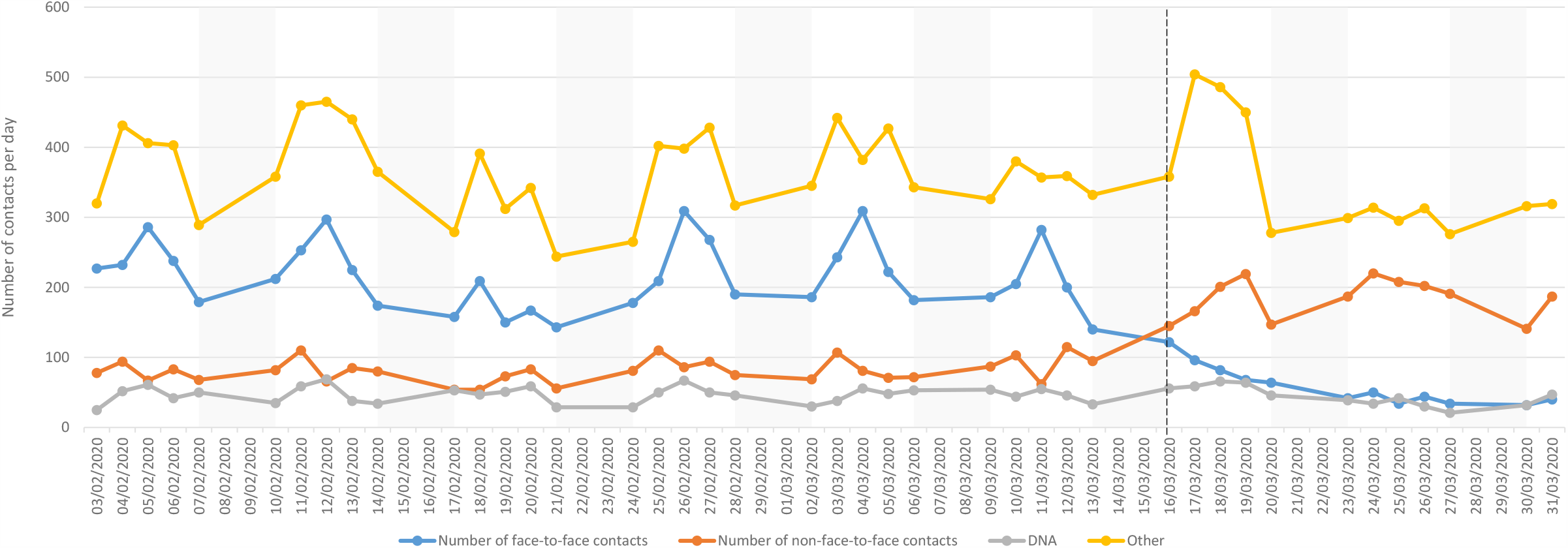
Child and adolescent Community Mental Health Team contacts by contact type (daily; Monday-Friday; February-March 2020)

**Figure 12:**
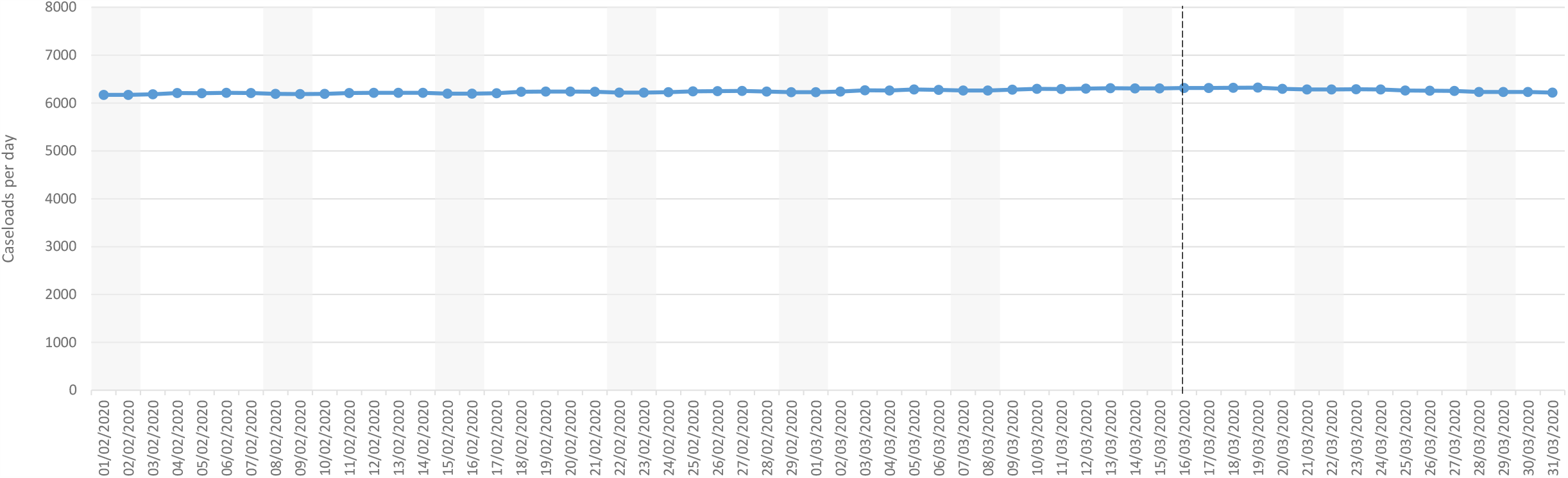
Child and adolescent Community Mental Health Team caseloads (daily; February-March 2020)

**Figure 13:**
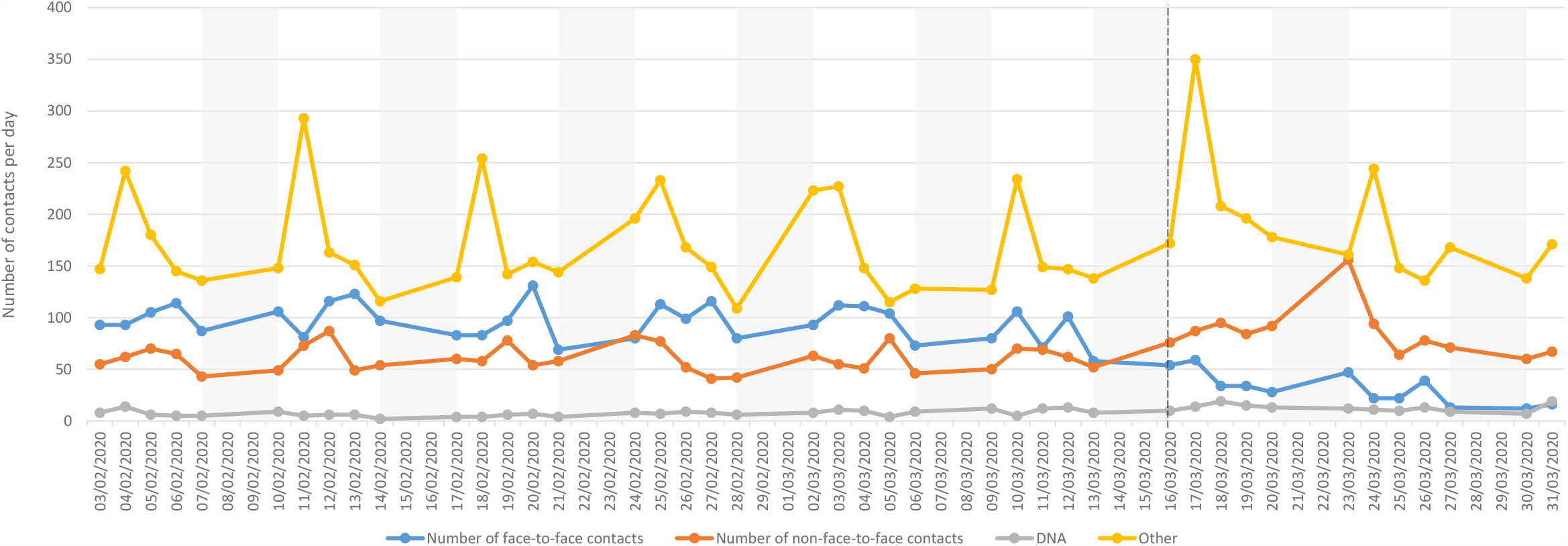
Older adult Community Mental Health Team contacts by contact type (daily; Monday-Friday; February-March 2020)

**Figure 14:**
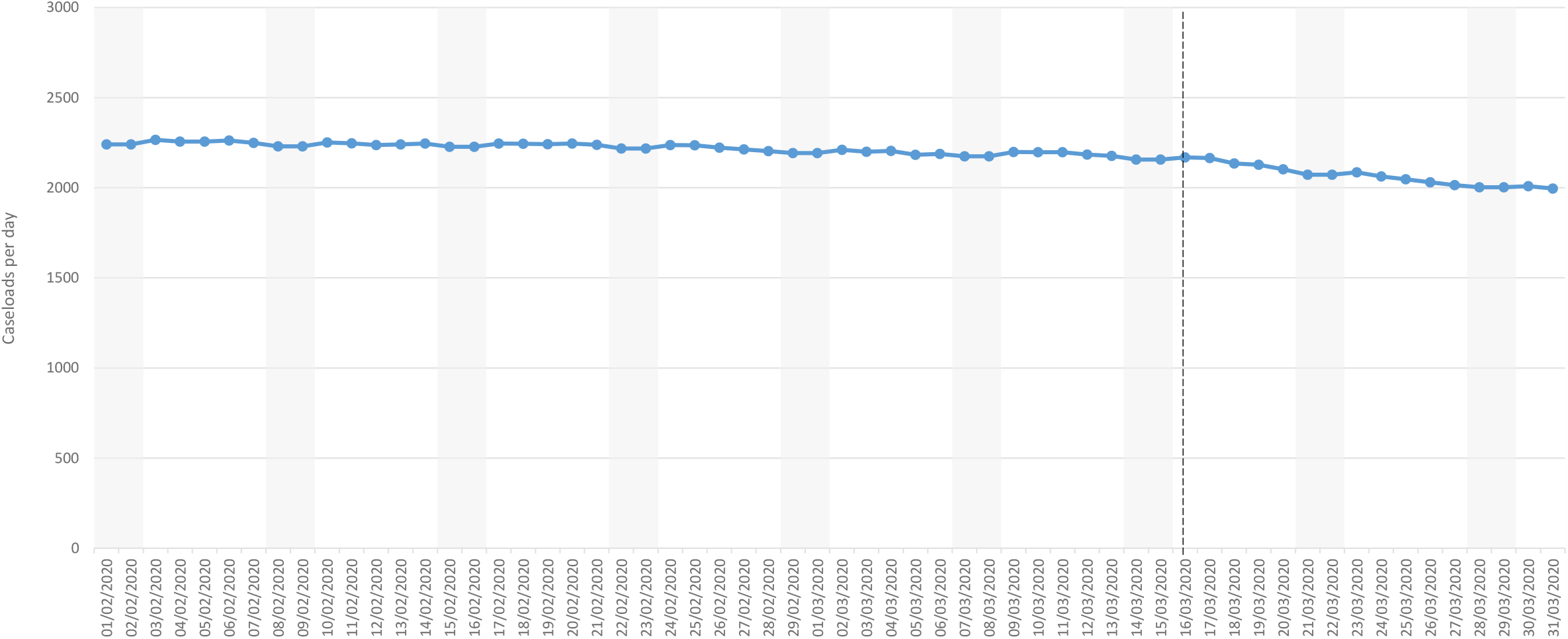
Older adult Community Mental Health Team caseloads (daily; February-March 2020)

**Figure 15:**
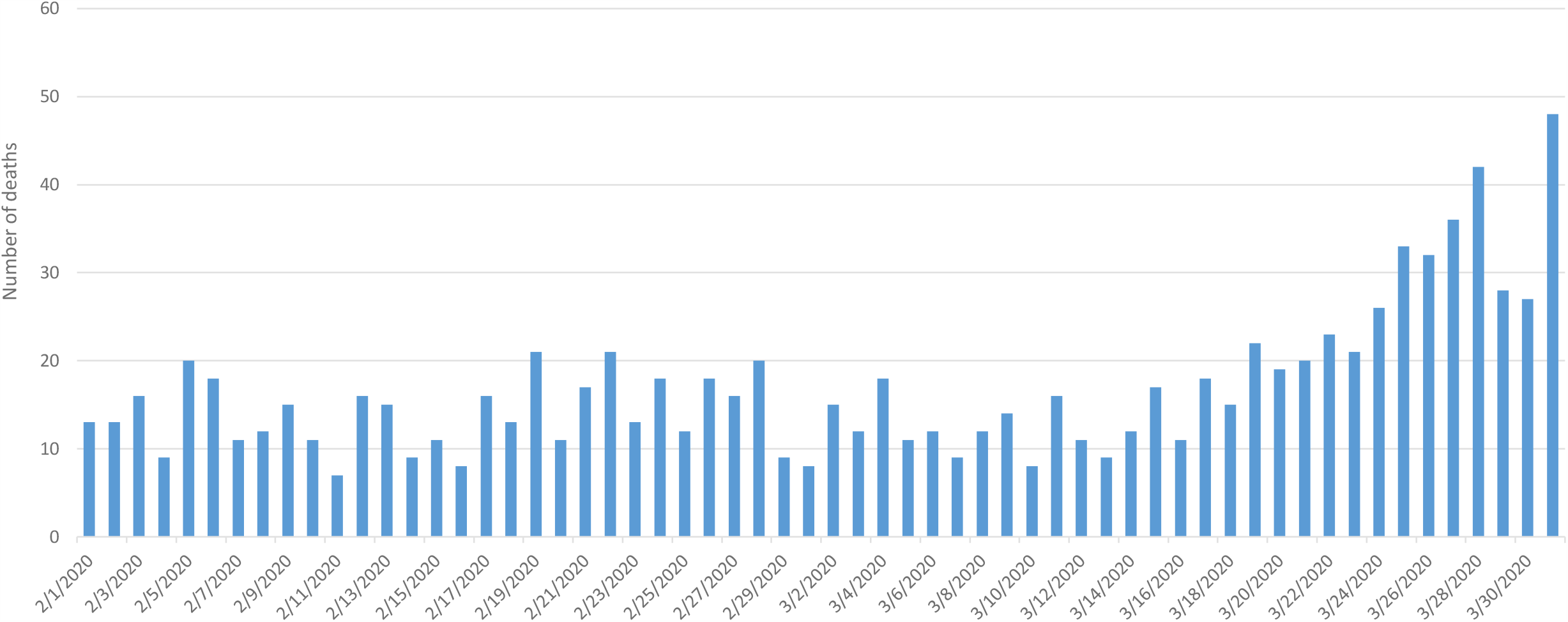
Total number of deaths for all previous SLaM service users (daily; February-March 2020)

## DISCUSSION

We present findings from an early extract of data from a large multi-team provider on changes in mental health service provision before and in the initial stages of the COVID-19 pandemic first wave in the UK, analysing these in relation to the enacting of a national social distancing policy. In summary, all grouped liaison, home treatment and community mental health services had substantially reduced face-to-face patient contacts, with variable and only partial compensatory increases in non-face-to-face contacts. Many had also reduced their caseloads, although those for working age CMHT and CAMH services remained level. Numbers of patients in inpatient care had also been substantially reduced. Daily numbers of deaths in past and present patients had increased significantly over a relatively short time period.

At the outset, the potential impact of the COVID-19 pandemic was widely discussed in a general sense, rightly focusing on the initial priorities of infection control, treatment options for severe complications, and the preparedness of critical care services (8). However, a second wave of evidence gathering grew in importance, because of the potentially sizeable indirect consequences on other healthcare sectors. For mental healthcare, there continues to be a need to understand the population-level impact of both viral infection (severe or otherwise) and the social distancing being imposed by many national governments (4;9;10). For people with pre-existing mental disorders, there has been a concern expressed that vulnerability to COVID-19 infection may be higher than expected – because of infection susceptibility, comorbidity and barriers to health service access (11). Also discussed has been a potentially higher risk of mental health deterioration due to the stress of the pandemic itself, the stress of quarantine as a consequence (12), and reduced access to routine outpatient visits for evaluations and prescriptions, and there were concerns raised about a higher risk of suicide as a result of the rapid social, economic and health changes (13). In addition, there may be higher rates of new presentations to services as a result of complex bereavements and post-traumatic stress disorder following severe hospitalised infections. Some of these outcomes may still only become apparent after enough follow-up has been accrued for adequately powered analyses, by which time it may be too late for intervention. At the time the data were extracted for this report, the scale of mental healthcare changes had not yet been fully quantified, although recommendations had been made in China for tighter admission criteria and reduced hospital outpatient visits, amongst others (14).

While it was not our intention in this paper to investigate factors underlying the observed service changes, many will be unsurprising. Clearly an inevitable outcome of social distancing, coupled with the rising awareness of staff risk from infections (and of the potential for staff-to-patient transmission), was a reduction in face-to-face clinical assessments. At the time of analyses, these had not been fully matched by increases in non-face-to-face assessments (for example, those carried out over the phone or via video calls) in all services evaluated. While it is possible that contacts labelled as ‘other’ might have included some direct assessments, numbers of contacts in this category were not markedly changed over the period of interest. The largest reductions after 16^th^ March in total contacts were seen in Liaison services, followed by HTT services and then CMHTs. On the other hand, reductions in median daily caseload were highest in older adult and working age HTT services followed by small reductions in Liaison and older adult CMHT caseloads and no significant reductions in working age CMHT and CAMH services.

A rise in daily numbers of deaths after 16^th^ March was also noted, although these findings were derived over a relatively short duration of surveillance. Considering contemporaneous national reports for total mortality (15), the 62% increase observed in the SLaM Case Register was of a similar order to the national 99% increase in care home residents, 72% increase in hospital inpatients and 51% increase in private home residents reported at the time (15).

Strengths of this study included the relatively ‘real time’ data from a large mental healthcare provider, which allowed investigation of very early changes in service activity following dramatic and rapid transitions. Clearly generalisability needs to be evaluated, as there may be a number of local and national factors that influence service transitions. London saw some of the earliest accelerations in COVID-19 infections in the UK and there will have been pressures arising from local medical services (e.g. to free up mental health inpatient beds for ‘overflow’ from acute care) which are likely to have been felt by other London mental healthcare providers, but which may have occurred ahead of other areas of the UK. Considering other limitations, data for this manuscript were drawn from specific services of interest and do not reflect SLaM’s full activity; they were also combined by broad service categories and we did not seek to investigate within-service variation. Daily contact numbers were quantified from structured fields applied to case note entries and might reflect recording behaviour rather than activity levels (e.g. if multiple contacts were recorded within one entry); also, the dichotomy between face-to-face and non-face-to-face contact is a relatively crude one and does not reflect the quality or depth of assessments being recorded. Finally, statistical power was limited because of the short period evaluated, as well as being limited by lack of data on cause of death and applied to a heterogeneous sample of past and present service users.

First author’s note, in the interests of transparency: the findings reported in this paper were submitted in manuscripts to *BMJ Open, BJPsych Open*, and *BJPsych Bulletin*, with the first submission on 21^st^ April 2020 after a data extraction on 2^nd^ April 2020 (daily deaths data were updated in a subsequent extraction for later submissions). The findings were adjudicated to be insufficiently informative by the first two journals and the manuscript was rejected by the third because of difficulties obtaining reviews. By the time final feedback was received (31^st^ July 2020), the study was judged by the authors to be too out of date for further attempts to seek peer-review. In the meantime, a number of approaches received from policy bodies for dissemination of findings reported here could not be accommodated because of results being under consideration for publication. The experience of attempting to follow the traditional academic publishing route in the context of rapidly changing circumstances requiring up-to-date data is a reason underlying recent dissemination of pandemic-relevant output from CRIS primarily via pre-print (16-19). The manuscript presented here has been amended from that originally submitted to remove statements of inference that might warrant peer-review.

## Data Availability

Data are available on request from the corresponding author.

